# Identifying SARS-CoV-2 regional introductions and transmission clusters in real time

**DOI:** 10.1101/2022.01.07.22268918

**Authors:** Jakob McBroome, Jennifer Martin, Adriano de Bernardi Schneider, Yatish Turakhia, Russell Corbett-Detig

## Abstract

The unprecedented SARS-CoV-2 global sequencing effort has suffered from an analytical bottleneck. Many existing methods for phylogenetic analysis are designed for sparse, static datasets and are too computationally expensive to apply to densely sampled, rapidly expanding datasets when results are needed immediately to inform public health action. For example, public health is often concerned with identifying clusters of closely related samples, but the sheer scale of the data prevents manual inspection and the current computational models are often too expensive in time and resources. Even when results are available, intuitive data exploration tools are of critical importance to effective public health interpretation and action. To help address this need, we present a phylogenetic summary statistic which quickly and efficiently identifies newly introduced strains in a region, resulting clusters of infected individuals, and their putative geographic origins. We show that this approach performs well on simulated data and is congruent with a more sophisticated analysis performed during the pandemic. We also introduce Cluster Tracker (https://clustertracker.gi.ucsc.edu/), a novel interactive web-based tool to facilitate effective and intuitive SARS-CoV-2 geographic data exploration and visualization. Cluster-Tracker is updated daily and automatically identifies and highlights groups of closely related SARS-CoV-2 infections resulting from inter-regional transmission across the United States, streamlining public health tracking of local viral diversity and emerging infection clusters. The combination of these open-source tools will empower detailed investigations of the geographic origins and spread of SARS-CoV-2 and other densely-sampled pathogens.

## Introduction

The massive scale of the SARS-CoV-2 sequencing effort has revealed deep inadequacies in our current methodology for phylogenetic analysis. Tools designed to work on small, sparse, static datasets have adapted poorly to the demands of a pandemic where tens of thousands of new genome sequences are generated and shared daily^7^. Some have made progress by adopting generalized statistical methods built for large data such as random forest regression^14^, but scalable phylogenetic solutions need to be developed. While our group, among others, has laid the groundwork for pandemic-scale phylogenetics ^3,12,13,20,23,25^, much remains to be done to translate evolutionary inferences to public health inference and response.

The unprecedented scale of the genomic sequencing effort requires novel approaches to evolutionary, medical, and public health inference. Some groups have developed phylogenetically informed statistics for identifying mutations associated with increased transmissibility and other fitness-related parameters^16,24^. In other cases, simple methods-such as the assaying of groups of identical samples-have been successfully applied to identify superspreader events and similar infection clusters^2,6^. Unfortunately, many analyses still lack scalable or phylogenetically informed approaches.

The intersection of geography and phylogenetics, phylogeography, has often relied on heavily downsampled and static trees or limiting their analysis to early stages of the pandemic^4,5,9,10,11,18^. While useful for assessing transmissions between countries and major introductions, downsampling limits our ability to assign specific samples to regional infection clusters or identify clusters of potential interest. Even with heavy downsampling, performing these analyses often required significant computational power, time and specialized resources^15^. Additionally, much of these analyses are not readily interpretable for an efficient public health response, lacking intuitive visualization and data exploration tools. There is therefore a significant need for fast, automated, scalable and interpretable phylogeographic approaches for an effective public health response to emerging situations.

To address this need, we present here a phylogenetically-informed summary heuristic (the “regional index”), implementation (matUtils introduce), and data exploration and visualization tool (Cluster Tracker: https://clustertracker.gi.ucsc.edu/) for identifying introduction events and associated clusters of descendants in a given region. Our approach can be used to efficiently identify infection clusters and evaluate transmission dynamics across dozens of regions and millions of samples. Results obtained using this method are congruent with widely applied analyses and are accurate when applied to simulated data. Our visualization platform enables researchers and public health workers to explore new SARS-CoV-2 introductions across the USA, updated daily with all available global public data. This work will empower real time research and public-health applications of genomic epidemiology during the SARS-CoV-2 pandemic and beyond.

## Results and Discussion

### Cluster Concept and Definitions

A cluster is a set of closely-related samples from the same region and descended from a common ancestor with a regional introduction event. In the phylogenetic tree, they appear as a set of leaves (samples) from a given geographic region that are descended from a shared common ancestor. A cluster may be monophyletic or paraphyletic, depending on whether some descendants of the cluster common ancestor left the geographic region. We consider location, or region, as a categorical state across the phylogenetic tree. A regional transmission event is where a child node is from a different region than the parent node. These patterns reflect cases of infected travelers moving between regions, followed by local transmission and eventual sampling of a number of descendent infections.

### A Heuristic for Identifying Introductions and Clusters

The core of our heuristic is the “regional index”, which is a weighted summary of the composition of descendants of a node of a phylogenetic tree, roughly corresponding to our intuition that the virus represented by that node was inside or outside a specific area. It is defined as

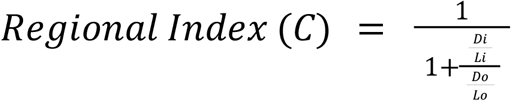

where “L_i_” is the number of downstream leaves that are in a given region, “D_i_” is the total branch length to the nearest leaf which is in that region, and “L_o_” and “D_o_” are the same for out-of-region leaves (Figure 1). On a tree inferred using maximum-parsimony, total branch length is equivalent to the distance in mutations between the query node and the descendant leaf.

**Figure 1:**
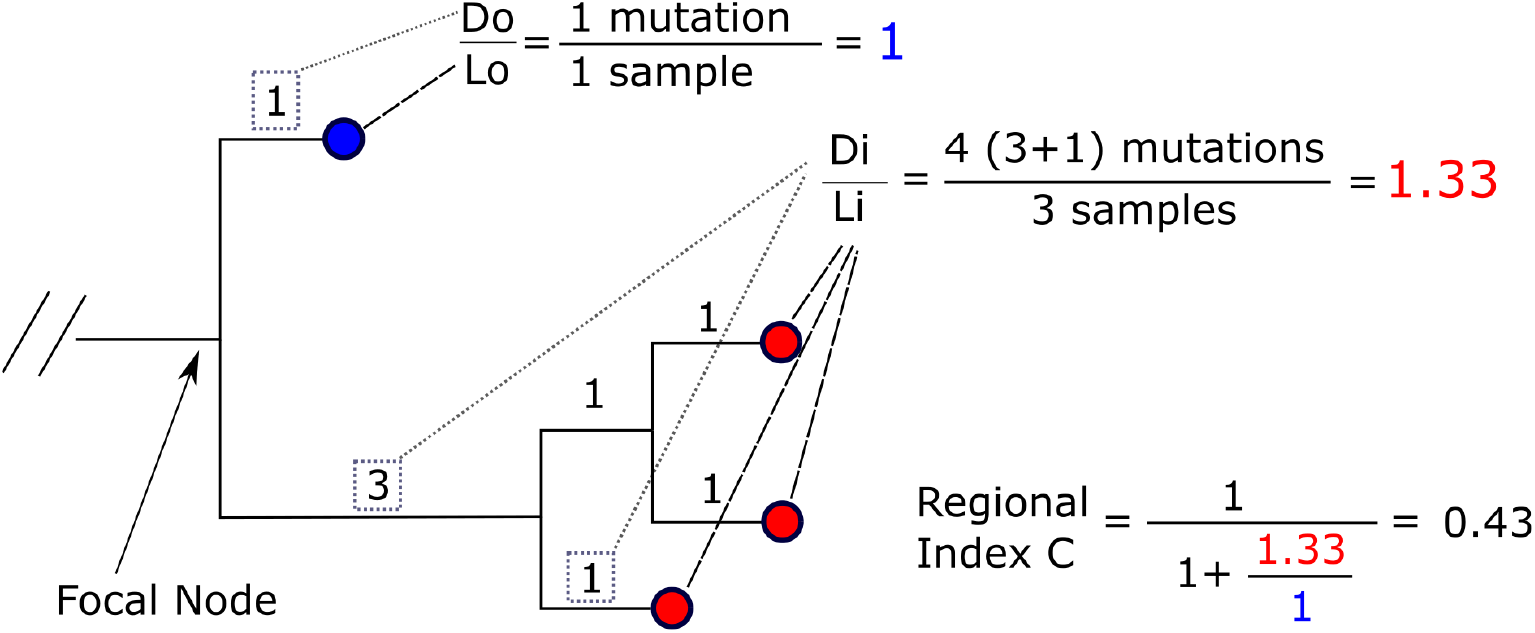
Example Index Calculation. The focal node at the base has an index value below 0.5, suggesting that it is out-of-region by our heuristic. Our introduction point is therefore along the long branch below the root, and the ancestor of the downstream in-region sample cluster would have existed along that branch.

We apply additional rules to handle cases where C is undefined. When a descendent leaf is genetically identical to the internal node and is in-region, C is 1. Similarly, when a genetically identical leaf is out-of-region, we treat C as 0. When such identical children exist both in and out of the region, we treat the node as in-region, as some infection with this genome must have existed in that region. We do not apply this index calculation to leaf nodes, which do not have children, and assume simply that the leaf is either in or out of the region as a given. This requires that each leaf included in the analysis be accompanied by accurate geographic location metadata.

This heuristic has several useful behaviors. For example, a sample identical to a specific internal node will always confer complete confidence about the location of that node, as we have sampled one genome that is identical to the ancestor directly. This can effectively identify nested clusters, where a new group of infections resulting from a regional introduction in turn produce clusters in other regions. It also accounts for the number of leaves downstream in our heuristic, on the assumption that introductions of a strain from one region to another require the lineage to be locally circulating in the origin region, but not necessarily lead to significant local transmission in the target region. This reduces the overall number of introductions we infer. Our heuristic strikes a balance between these two principles, allowing us to efficiently analyze a large phylogenetic tree with minimal metadata.

Once indices for a given region have been calculated for each node, the second step is to identify clusters of samples putatively associated with an introduction. This is accomplished on a per-sample basis. The path from the sample to root is traversed and the indices for each ancestor being in the focal region is noted. Generally, the index declines from 1 to 0 along the ancestry path from leaf to root. The introduction point is called where the index for an ancestor being in-region is below 0.5, or the root, whichever is encountered first.

As this heuristic is independent and specific to a region, it can be computed for an arbitrary number of regions across a single tree in parallel. When multiple regions are included, origins of putative clusters can be identified after introduction points are found by examining index scores across all other regions for the origin node and noting the region with the highest index. This metric can be calculated for one region of any size in a single post-order traversal with dynamic programming (see Methods), which makes it very fast to compute even on extremely large phylogenies with expansive regions.

### Evaluation of Our Heuristic Method

Our implementation is part of the matUtils online phylogenetics package^13^ and uses the efficient mutation annotated tree protocol buffer format and associated library^23^. To test runtime efficiency conditioned on a tree, we applied random subsampling and recorded time to compute our heuristic for a single region. We found that it takes less than forty five seconds on a single thread even for trees of more than two million samples (Supplementary Table 2).

To validate our results, we performed simulations consistent with viral evolutionary dynamics with inter-region dispersal events using the tools VGSim^20^ and phastSim^12^ (see Methods). We found that our heuristic with default parameters recovered the true geographic location of internal nodes up to 99.8% of the time under realistic conditions for SARS-CoV-2 across an exactly correct bifurcating tree. We further attempted to model our ability to correctly recover clusters on a simulated tree with collapsed branches and realistic mutation rates for SARS-CoV-2. In comparing the clusters we recovered with the true set, we obtained an adjusted Rand index of up to 0.999. This suggests that our approach is generally quite accurate, though high migration rates or extremely low mutation rates can be confounding, as these scenarios are associated with minimal geographic and phylogenetic signal respectively (Supplementary Table 1; See Methods). More practically, this implies that our method will perform best when within-region transmission is substantially more common than between-region transmission (as in *e.g*., country-level or state-level analyses).

To compare our results to widely used but much slower (days to months) analyses, we used our method to replicate a published phylogeographic analysis for the SARS-CoV-2 pandemic. Alpert et al^1^ used Bayesian phylogeography to identify 23 distinct introductions of B.1.1.7 into the United States as of March 4th 2020. We obtained their subsampled tree and applied our heuristic using country labels to define the relevant regions (see Methods). With our method, we exactly replicated their identified clusters (Adjusted Rand Index 1.0). Alpert et al^1^ additionally predicted “sink” states, or the state to which each of the 23 introductions initially transmitted. We find that for all 23 clusters, samples in the identified sink state are closest or tied for closest in branch length to our inferred introduction point. This suggests that our approach can produce results congruent with more complex statistical models in a fraction of the time.

### Global SARS-CoV-2 Transmission Dynamics and Infection Clusters

Using our method, we traced transmission clusters in 102 countries from across the world (Figure 2A) using the global parsimony phylogenetic tree, built from 5,563,847 available sequences on GISAID^21^, GenBank^19^, and COG-UK^25^ on 11-28-2021 (see Methods). Cluster size is highly skewed (Figure 2C), with approximately 20% of distinct regional clusters containing 89% of samples. This suggests that the majority of novel introductions do not lead to the establishment of a new locally-circulating strain, consistent with previous findings^5^.

**Figure 2:**
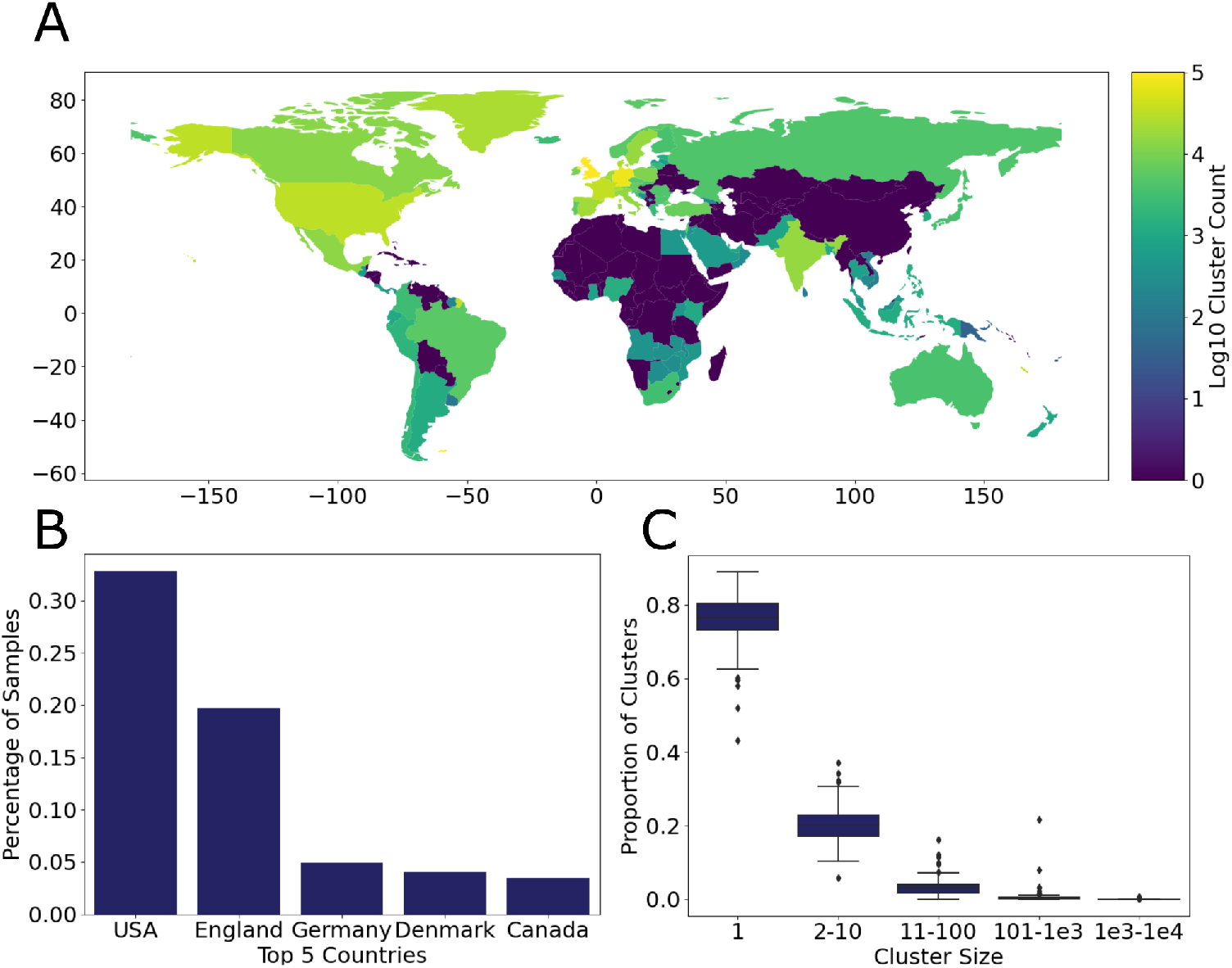
Global Distribution of SARS-CoV-2 Transmission Clusters. **A:** The log count of clusters detected across each of the 102 countries surveyed. The number of clusters detected is largely a function of total local sequencing effort. **B:** The five countries with the highest representation in the data. The USA and England together constitute more than half of all available sequences. **C:** Cluster sizes are consistent across countries. Most clusters are small, implying most newly introduced SARS-CoV-2 lineages quickly die out.

Global contributions to sequence repositories are notably biased, with 51% of all samples belonging to either the USA or the United Kingdom (Figure 2B). This is a significant restriction on global transmission analysis, especially as the inference of the origin of a cluster is highly dependent on robust sequencing at the origin (see Methods). We therefore narrowed the next step of our analysis to the United States, which has robust and relatively comprehensive sequencing across each state as well as detailed state-level metadata for the vast majority of available samples.

### SARS-CoV-2 Transmission Into and Across the USA

We identified more than three hundred thousand distinct state-level SAR-CoV-2 infection clusters in the United States over the course of the pandemic, as of November 2021 (Figure 3). Approximately 84% of these clusters have an assigned origin using our method (see Methods). Only 7% of our clusters appear to be of international origin, with the majority reflecting transmission within the USA. Mexico and Canada are among the most common international origin regions, in line with expectations given their long land borders (Supplementary Table 3). England is also relatively common, likely because it is very well sampled. This indicates that it is possible that some clusters originate from less sampled intermediate regions and are assigned to the UK or other highly sampled locations. This suggests that relative sequencing effort in a given region is an important bias with respect to accurately identifying the origins of newly identified clusters and results should be interpreted with caution. International introductions rates are correlated with higher total sampling and therefore population size, particularly for California, Texas, New York, Massachusetts, and Florida (Figure 3B).

**Figure 3:**
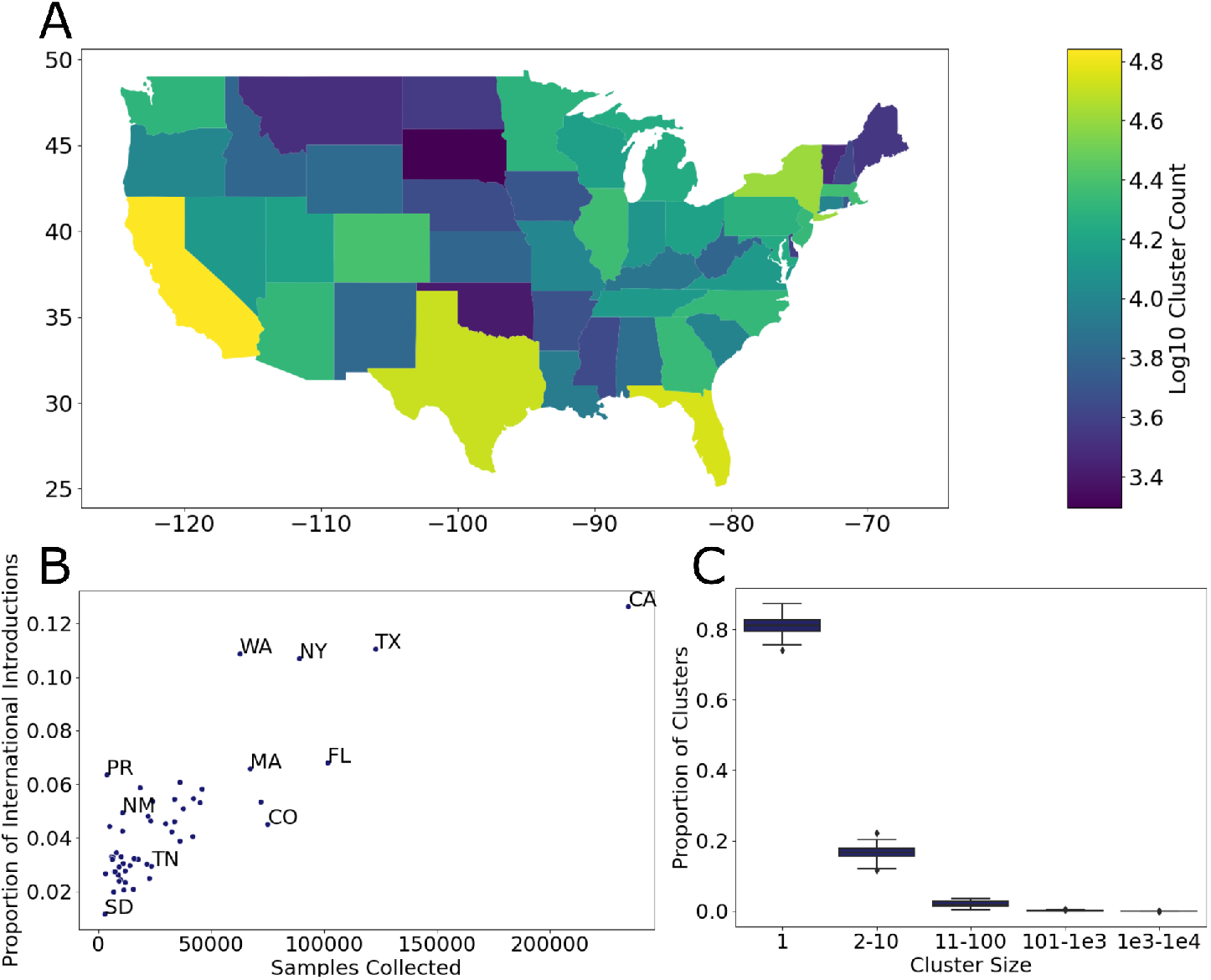
International and Interstate Introductions across the USA. **A:** The log count of clusters identified across the continental USA. California, Texas, Florida, and New York are associated with the greatest number of unique clusters. **B:** The proportion of international introductions in each state plotted against total samples collected in that state. This relationship is largely linear, reflecting the correlation between sampling, population size, and levels of international travel. PR (Puerto Rico) exhibits relatively more international introductions for its sampling than other territories and states of the United States. **C:** The distribution of cluster sizes across states. These are largely consistent with clusters identified at the international level.

Within the USA, introductions come from a mix of neighboring states and high-population travel centers (Supplementary Table 3). We attempt to mitigate sampling biases-resulting from larger populations, higher case rates, increased sequencing, or other factors that are not specific to geography-by calculating a log-fold enrichment for rates of introduction from a given source region (see Methods; Figure 4). Note that while log-fold enrichment may reveal spatial relationships, it does not reflect the absolute importance of a region as a source or sink of viral transmission.

**Figure 4:**
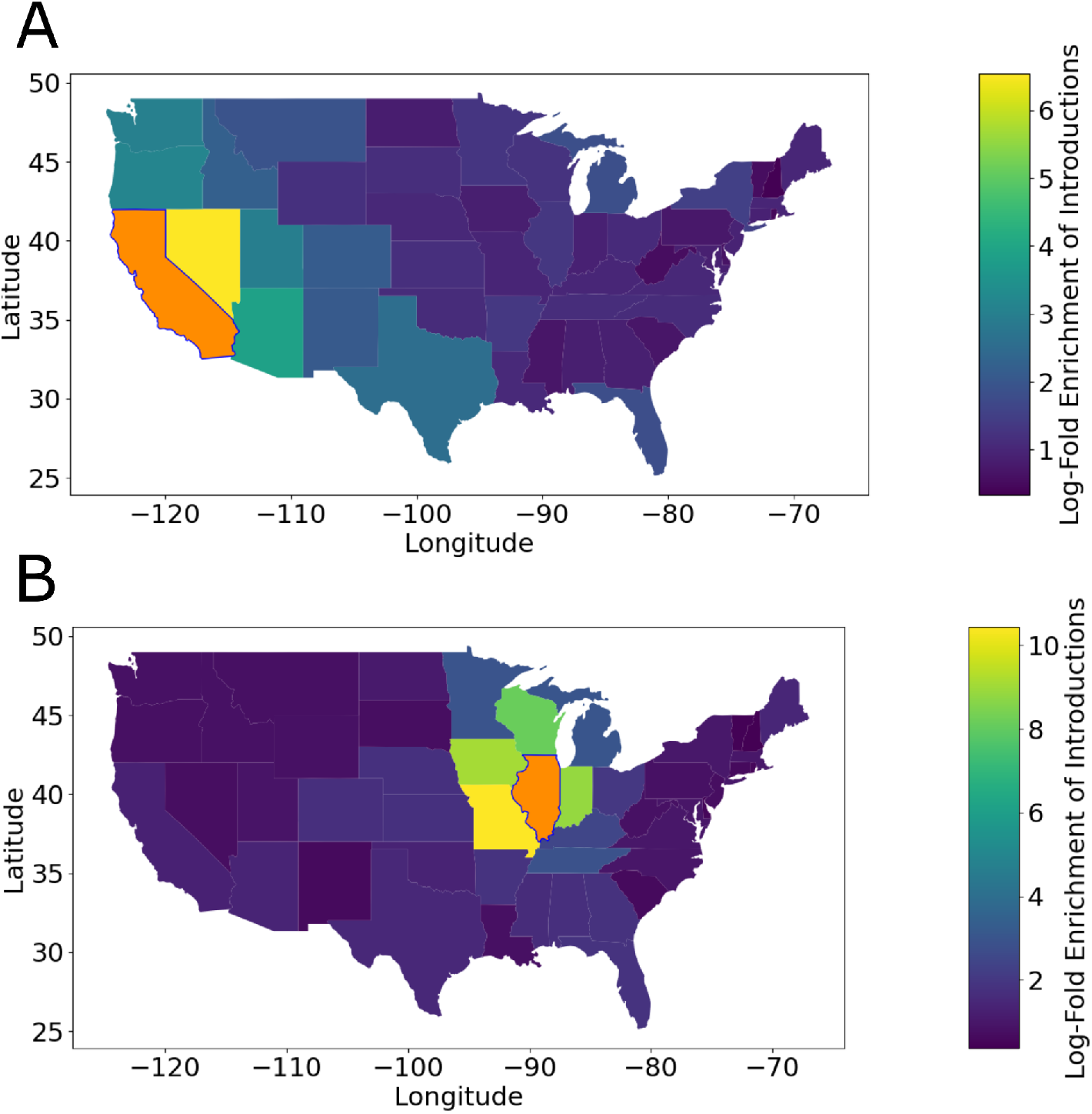
Log-Fold Interstate Transmission. **A:** Interstate introductions of COVID-19 into California are relatively more likely to originate on the West Coast, particularly from Nevada. **B:** Interstate introductions of COVID-19 into Illinois are relatively more likely to come from the immediate surroundings, particularly Iowa and Missouri.

As with results from international introductions, we also find an enrichment for introductions that originate in geographically adjacent states. Log-fold enrichment is more than five times greater for neighboring states than for non-neighboring states within the USA (p=1.5e-117, Mann-Whitney U). Simple counts of inferred introductions are also enriched to a lesser extent between geographically adjacent states (p=2.2e-16, Mann-Whitney U). This suggests that SARS-CoV-2 transmission over interstate land borders is a major mechanism for spread within the USA. These results are largely in line with previous results in other viruses^8^ and SARS-CoV-2^22^, suggesting that this heuristic is capturing and summarizing true geographic structure within the global SARS-CoV-2 phylogenetic tree.

### A Daily-Updated Website To Explore SARS-CoV-2 Clusters in the USA

To make the results of this work broadly useful for the research and public health community, we have developed a visualization and exploration platform. Cluster-Tracker is a publicly-available, daily-updated website displaying the latest results for applying our heuristic to sequences collected from across the United States of America interactively (clustertracker.gi.ucsc.edu; see Methods; Figure 5). Cluster-Tracker is open-source with a flexible backend pipeline that allows any user to construct a similar site for any set of regions they have geographic information and sample identification for (https://github.com/jmcbroome/introduction-website).

**Figure 5:**
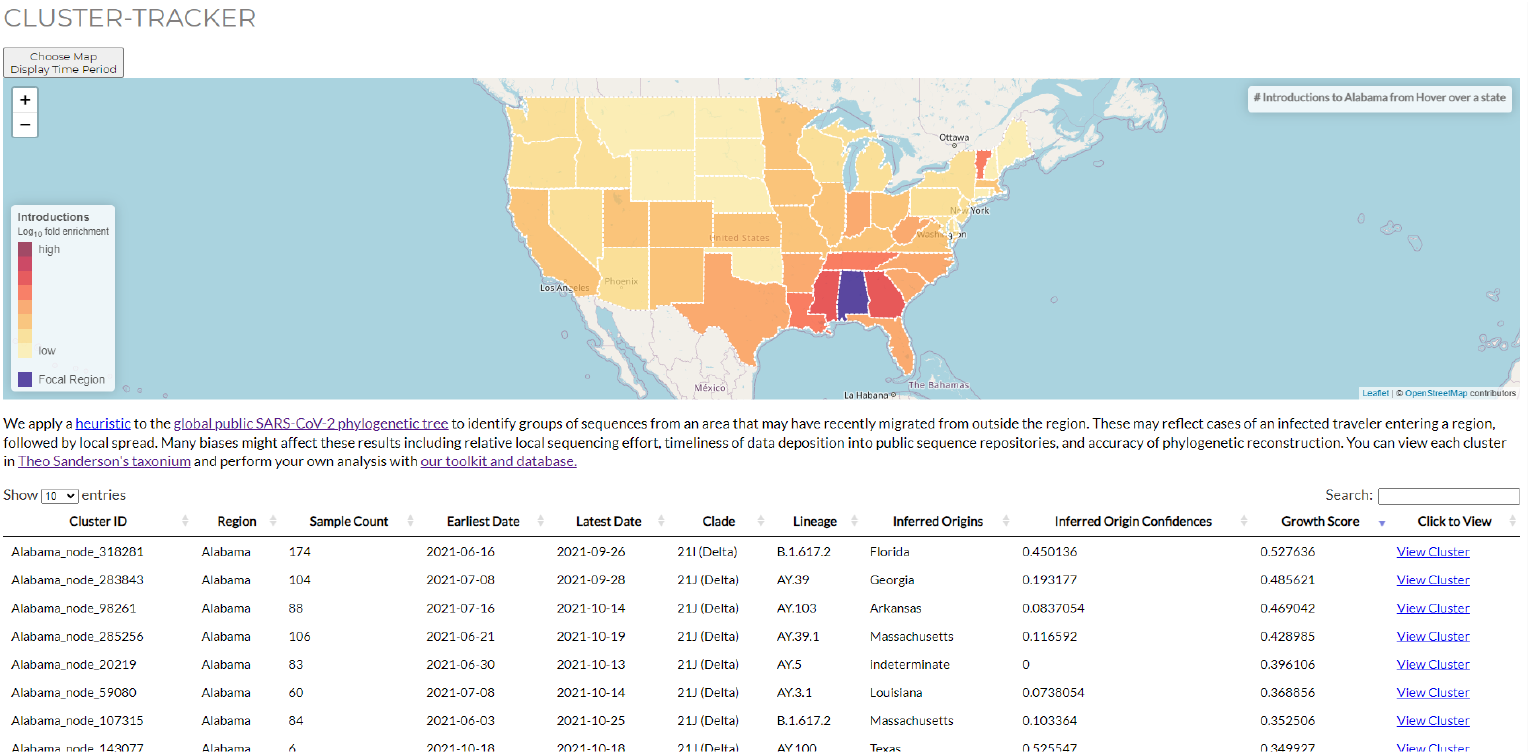
the Cluster-Tracker site. The Cluster-Tracker tool is updated daily at clustertracker.gi.ucsc.edu. Users can interactively explore the latest results of our heuristic applied to each of the continental United States, by sorting the interactive table, selecting states to focus on in the map, and using the Taxonium tree-viewing platform to examine clusters of interest in detail.

Cluster Tracker is composed of two primary sections and some descriptive text (Figure 5). The first section is an interactive map of the United States. In the default view, this map is colored by the number of clusters detected across each state throughout the course of the pandemic. The true number of introductions into a given region is likely to be substantially larger because many small clusters will not be sampled by ongoing viral surveillance efforts, but major local transmission clusters should be represented. By clicking on a state, the site changes to a view specific to that state. In this view, the map is colored by the log-fold enrichment of introductions from each other state to that state.

The second section is a sortable, searchable table display of the highest priority clusters. In the default view, these are the top 100 clusters overall as sorted by “growth score”. We define “growth score” as the square root of the number of samples divided by the number of weeks since the introduction occurred. The goal of this metric is to weight clusters by relative size and how recently they entered a given area, so that clusters of interest to public health appear first. When a state is selected, this table changes to the top 100 clusters obtained from that particular state. Basic information including clade, lineage, the earliest and latest dates of detection, and inferred origins are displayed for each cluster. The “inferred origin confidences” column is the highest or tied for highest regional index among all other regions for the parent node to the cluster origin, with a floor of 0.05 below which the cluster is simply marked “indeterminate”. The “inferred origins” column is the regions which match these scores, and generally represents our best guess at the origin of this cluster. The last column of the table contains links to the Taxonium viewer (https://github.com/theosanderson/taxonium) which will automatically render the full tree and zoom to the cluster of interest when opened (Figure 6). Full results and the taxonium protocol buffer file, which encodes the tree and all cluster IDs, are available to be downloaded at the bottom of the page.

**Figure 6:**
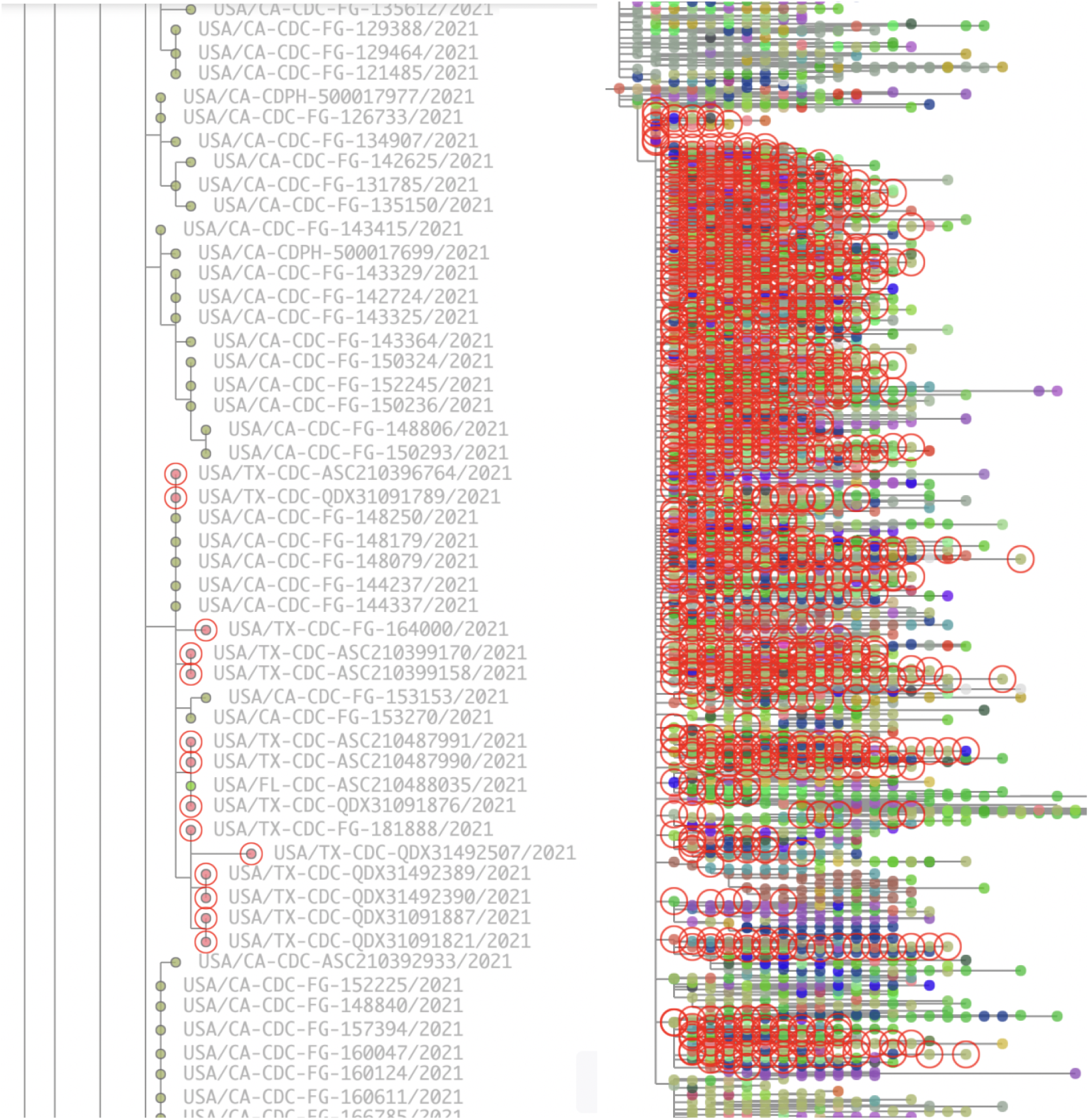
Example clusters in the taxonium viewer. (A) An example cluster in Texas (circled in red) that is inferred to have originated from California (Regional Index = 0.94). There are many samples from California closely related to the cluster’s common ancestor, supporting California as the most likely origin. (B) A different, much larger, 9,533 leaf cluster in California. This represents a lineage of SARS-CoV-2 commonly circulating in California, descended from one of the original introductions of the Delta variant into California in mid June 2021. Descendants from this cluster have transmitted to other regions many times, but members of this cluster have been found in California as recently as December 7th 2021.

The goal of this resource is to make cluster identification, exploration, and prioritization more accessible and digestible for public health offices and policy makers. A significant roadblock for public health action is the sheer quantity of daily new data and the speed with which we can draw inferences from these data. Cluster-Tracker can assist exploration and prioritization of the latest genome sequences, quickly identifying the clusters most likely to be of interest for public health action for a given region. Our construction pipeline is flexible and can be applied for any set of regions (*e.g*., county-level), allowing groups anywhere to construct web interfaces for intuitive SARS-CoV-2 phylogenetic data exploration.

### Conclusion

The pandemic has made the need for rapid and powerful tools to unlock the potential of pandemic-scale genomic epidemiology. The method we developed and the efficient software package we provide will empower researchers worldwide to make fast inferences from vast sequence datasets. Our results have revealed geographic structure at scales below the level of pango-lineage^14^ within the global SARS-CoV-2 phylogeny. We have provided tools and resources with which to explore this geographic structure and draw useful inferences for specific areas. Additionally, to empower public health officers and the public to explore the spread of SARS-CoV-2 across the USA, we developed an accessible open-source interactive interface for our results, which can automatically compute and display introductions and clusters with each update to the global phylogenetic tree. Our work can support public health groups across the world to quickly understand and apply insights obtained from the latest genomic data.

## Methods

### matUtils Implementation

We implemented a calculation of this heuristic as a part of our online phylogenetics package, matUtils, under the command “matUtils introduce”^13^ (https://github.com/yatisht/usher). Our implementation uses dynamic programming based on a post-order traversal to compute the regional index for each node in the tree in a single pass for each region (equation 1). This is because the four parameters which define regional index-distance to the nearest descendent and total descendents for in-region and out-of-region-can be computed from these same metrics for each child of a node plus the branch length to each child. The total number of leaves descended from a query parent node is the sum of all leaves descended from each of their children, and the shortest distance traversed to a leaf is the minimum of each child’s minimum distance traversed plus the branch length between that child and the query parent. Therefore, by computing it first for nodes with only leaf children, then progressively deeper internal nodes, we only have to reference the children of each internal node and check their stored values instead of having to traverse from each node. This step is optionally parallelized across distinct regions, if multiple regions are passed.

The secondary step is an ancestry traversal for each sample in the tree, identifying the most recent ancestor which has a regional index below the set threshold, which is inferred to be the introduction point for this lineage. Once introduction points have been inferred for each sample, samples are grouped by shared introduction points into clusters, basic statistics and information are computed, and results are reported.

Ultimately, our implementation can compute this heuristic, identify clusters, and report all results in less than two minutes for a tree containing more than two and a half million samples (Supplementary Table 2). The speed of calculation is a major attraction of this heuristic approach over more complex Bayesian models. Calculating in minutes on minimal computing resources makes this method accessible and applicable to update results daily, identifying clusters and introductions as they occur and new data is uploaded globally. Accordingly, this implementation underlies our website Cluster-Tracker, which is updated with all new uploaded data each day and a recalculation of our heuristic.

### Handling Nested Clusters and Unstructured Regions

We implemented a few additional parameters that can be used to control behavior at the secondary cluster identification step. Once that is useful is setting a short-range maximum index requirement-that is, looking ahead at some additional number of ancestors and ensuring that each of those have a lower regional index than the intended ancestor node. Setting this parameter causes small nested clusters to be merged into larger overarching clusters. Another useful parameter is a minimum required branch length between the ancestor inferred to be in-region to its parent; if the branch length is less than the minimum, then the parent instead of the in-region node is inferred as the introduction point. Setting this parameter allows sibling clusters to be merged if both of their branch lengths are below minimum; this also resolves unstructured parts of the tree where large polytomies of identical samples with branch length 0 both in and out of a region are included.

### Prioritization and Bias Handling

Another significant point of consideration is cluster prioritization. This cluster identification method is based solely on the phylogenetic tree and simple sample-region association, and while this makes it lightweight and flexible, identifying clusters which died out locally months ago is not of use to public health offices doing real-time transmission cluster tracking. We therefore in our implementation sort the output by a “growth score”, defined as the square root of the number of samples associated with the cluster divided by the time in weeks from the oldest sample in the cluster to the current date plus one. This means that large, recent clusters will appear at the top of any output tables, and makes the method more easily accessible when thousands of clusters are being inferred simultaneously.

When using this method to examine inter-region transmission dynamics, we rely on comparable and significant levels of sequencing in order to identify introduction origins. Intuitively, the less sequencing is performed in a region, the less likely we are to recognize sequences from that region when they appear in another region. We can compensate for this bias to an extent by calculating log-fold enrichment of introductions between regions. This is computed as

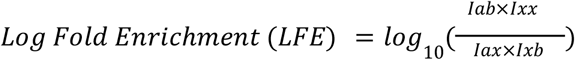

Where I_ab_ is introductions from region A to region B, I_xx_ is introductions from any region to any region, I_ax_ is introductions from region A to any other region, and I_xb_ is introductions from anywhere to region B. This computation can remove biases in rates of detected introduction which would apply to any pair of regions, but requires many regions to be computed as points of comparison. This score is used to color the map on Cluster-Tracker when a state is selected and has a very strong correlation with geographic distance.

### Simulation for Validation

To assay the performance of our heuristic, we fully simulated a pandemic phylogeny with VGsim^20^ and phastSim^12^. From the resulting mutation-annotated tree, we calculated true node region states based on VGsim’s migration event output and applied our heuristic with matUtils^13^. We then computed accuracy as the proportion of internal nodes which have a heuristic value above 0.5 for the true state. Leaves are excluded from this calculation as they are taken as an input in our heuristic and will always be 100% accurate.

For our specific results, we simulated a one-million-leaf SARS-CoV-2 tree under a simple model with two equivalently-sized regions with an even rate of migration between them, no strain or site selection and complete immunity for recovered individuals (Supplementary Table 1). We included a lockdown parameter starting at 5% infected and ending at 1% infected, with a 10-fold reduction in transmissivity under lockdown, and a sampling multiplier of 0.2 in order to deepen the tree by effectively extending the time for one million samples to be collected.

ARI (Adjusted Rand Index) and IAC (Internal Assignments Correct) are our quality metrics. ARI represents how well our method correctly groups samples into true clusters descended from a single introduction event. ARI performs best when migration is low, leading to large and clean clusters which are easily separated heuristically, and performs somewhat better when scale is increased. IAC is the proportion of internal nodes which are assigned to the true region by our heuristic across the bifurcating tree. It is computed on the correct bifurcating tree because collapsing true nodes from different regions leads to nodes that are naturally indeterminate. IAC is generally robust, only performing slightly worse with an increased migration rate, likely as deeply set internal nodes tend towards indeterminacy with high distances to many leaves across different regions. This suggests that the primarily limitation of our heuristic is simply the number of mutations available to distinguish samples from across varying regions rather than any structural or fundamental issues.

All code for this simulation is available as a modular and reproducible Snakemake pipeline at github.com/jmcbroome/pandemic-simulator.

### Global Phylogenetic Tree Construction

At UCSC we maintain a large phylogeny of all GISAID^21^, GenBank^19^, and COG-UK^25^ sequences using the script https://github.com/ucscGenomeBrowser/kent/blob/master/src/hg/utils/otto/sarscov2phylo/update Public.sh and the UShER online phylogenetics suite^12,21^. Updates are performed daily by obtaining all newly uploaded sequences from each database and placing them on the previous day’s global phylogenetic tree with UShER (see McBroome et al)^13^.Starting with our phylogeny updated on 11-28-2021, we pruned all samples with long branch lengths and path lengths using the matUtils parameters *--max-branch-length 45* and *--max-path-length 100* and performed a round of optimization with an SPR radius of 8. The resulting phylogeny contained 5563847 samples with a total tree parsimony of 4847954.

### Computing USA state transmission

We obtained the latest mutation-annotated phylogenetic tree representing the entirety of all public samples and all samples available on GISAID on 11-28-2021. As the standard format for publicly uploaded SARS-CoV-2 sequence identifiers is “Country/(Area)-CollectingAgencyInfo/Year|Date”, we extracted sample labels for samples in the USA by identifying samples with names beginning with “USA/” and then extracting the two-letter state code, if it matches with a two-state letter code. This resulted in 1764019 labeled samples belonging to the USA. Samples from outside the USA were labeled by country; countries and ambiguous labels with less than 500 samples in GISAID and public data were excluded and their samples removed. Samples from “mink” were additionally excluded as they may not be from human sources. The resulting tree contained 5237796 of the total of 5563847 samples available, reflecting more than 94% of all SARS-CoV-2 genomic data collected and incorporated to date.

We applied matUtils introduce with default parameters to this tree and sample set and produced the full by-sample output. After computing basic statistics, we calculated log-fold enrichment of introductions between all pairs of states, and a selection of other countries to and from the USA. All code for this paper is provided at https://github.com/jmcbroome/cluster-heuristic).

### Cluster-Tracker Website Development

All relevant javascript and some example data files are provided at (https://github.com/jmcbroome/introduction-website). This github includes a brief description of how to set up a local test site and run the backend pipeline for generating new results to display for your regions of interest. It is based on Leaflet (https://leafletjs.com/) and DataTables (https://datatables.net/) for the primary view, and includes links to the Taxonium tree viewer (https://taxonium.org/) for detailed cluster exploration.

We include Python scripts to create the backend data for the website display, contained in the “data” directory. This includes two versions of the primary pipeline, one specific to the United States which fills in many default parameters and uses data included in the repository, and one more flexible pipeline which given a tree, labels, and a geojson can create an equivalent website for any set of regions.

### Comparison with Published Studies

To compare our approach to that of Alpert et al 2021^1^, we retrieved the Auspice JSON they used to generate Figure 3 from (https://github.com/grubaughlab/CT-SARS-CoV-2) and obtained table S3 from their supplementary data online, which contains cluster labelings for samples from the tree represented by the JSON. We converted the Auspice JSON to the UShER MAT protobuf format using python. We labeled all samples in the resulting tree by their country of origin and ran matUtils introduce with default parameters. The resulting labels were compared to the cluster labels presented in table S3 and the Adjusted Rand Index was computed across all labeled samples with scikit-learn^16^. We performed this analysis twice-once including all samples in their tree from any region and once excluding samples from the USA in their tree that were excluded from their clusters. The first method resulted in an ARI of 0.9 and the second a perfect 1.0; this discrepancy results from a single difference where a pair of large clusters, sibling to one another, are merged by our results when samples excluded from their clusters are included in our analysis. This is because a sample identical to the parent node of these two sibling clusters from the USA is excluded from Alpert et al’s ^1^ clusters. In any case, the clusters we identify are highly concordant with Alpert et al’s^1^ results. All code for this analysis is available on (https://github.com/jmcbroome/cluster-heuristic).

## Data Availability

All data was obtained from the public repositories GISAID, COG-UK, and GenBank. Full sample credit is included in Supplementary Data 1.
Code to replicate our analysis is available at https://github.com/jmcbroome/cluster-heuristic. Code for complete simulation of covid-like phylogenetic trees is available at https://github.com/jmcbroome/pandemic-simulator
Our implementation of our heuristic is implemented as part of matUtils https://github.com/yatisht/usher with additional documentation at https://usher-wiki.readthedocs.io/en/latest/
Our website source code is available at https://github.com/jmcbroome/introduction-website.

## Acknowledgements

We gratefully acknowledge helpful comments and feedback from Gage Moreno, as well as the submitting laboratories where genome data were generated and shared via GISAID^21^, COG-UK^26^, and GenBank^19^ and authors from the laboratories responsible for obtaining the specimens. Full sample credit is available in Supplementary Data 1.

## Author Contributions

JMB and RBC conceived and designed this research. JMB developed the heuristic, wrote the software, and performed comparison and validation experiments. JMB developed the website. JM contributed to the website and documentation for the website. JMB, RBC, and ABS edited the manuscript.

## Data Availability

Code to replicate our analysis is available at https://github.com/jmcbroome/cluster-heuristic.

Code for complete simulation of covid-like phylogenetic trees is available at https://github.com/jmcbroome/pandemic-simulator

Our implementation of our heuristic is implemented as part of matUtils https://github.com/yatisht/usher with additional documentation at https://usher-wiki.readthedocs.io/en/latest/

Our website source code is available at https://github.com/jmcbroome/introduction-website.

