## Supplementary Text and Tables 1-2 for "Identifying SARS-CoV-2 regional introductions and transmission clusters in real time"

**Supplement**

https://github.com/jmcbroome/cluster-heuristic/blob/main/supplementary_data_1.tar

**Supplementary Data 1:** Public data repository sample information for full accreditation.

| Scale | MigRate | NodesCollapsed | MutationsPerNode | Parsimony | ARI | TreeDepth | IAC |
|---|---|---|---|---|---|---|---|
| 0.001 | 0.001 | 569615 | 1.238430619 | 2476860 | 0.9372738283 | 19.847591 | 0.9984239984 |
| 0.001 | 0.005 | 589133 | 1.158238079 | 2316475 | 0.9322724852 | 13.792404 | 0.9915379915 |
| 0.001 | 0.01 | 580482 | 1.196507098 | 2393013 | 0.8889069058 | 14.157408 | 0.9844449844 |
| 0.001 | 0.05 | 583369 | 1.1999291 | 2399857 | 0.1462308541 | 17.652355 | 0.9430809431 |
| 0.005 | 0.001 | 209173 | 6.083390042 | 12166774 | 0.995476981 | 26.863182 | 0.9985989986 |
| 0.005 | 0.005 | 212422 | 5.985278993 | 11970552 | 0.9477928803 | 28.763355 | 0.9934049934 |
| 0.005 | 0.01 | 220101 | 5.747451374 | 11494897 | 0.914332382 | 25.352871 | 0.9871239871 |
| 0.005 | 0.05 | 204296 | 6.373095687 | 12746185 | 0.4702043148 | 28.995562 | 0.9503849504 |
| 0.01 | 0.001 | 116730 | 12.2084966 | 24416981 | 0.998693832 | 28.543488 | 0.9985619986 |
| 0.01 | 0.005 | 123025 | 11.37735469 | 22754698 | 0.9503027402 | 27.531197 | 0.9936439936 |
| 0.01 | 0.01 | 111625 | 12.78072189 | 25561431 | 0.4297117256 | 30.548616 | 0.9885259885 |
| 0.01 | 0.05 | 118963 | 12.08342404 | 24166836 | 0.4277722582 | 28.472488 | 0.9517919518 |

**Supplementary Table 1:** Results from a set of simulations generated via PhastSim and VGsim (see Methods). For reference, the real tree that we considered in this work had an overall parsimony of 4847954 and a mean tree depth of 35.

"Scale" is the parameter passed to phastSim --scale, representing a scalar applied to the branch lengths to rescale. Smaller values of scale imply fewer mutations per site per branch. "MigRate" is a reciprocal value representing the rate of migration events between two equally-sized "regions" under simulation. "Nodes collapsed" is the number of nodes which have no mutations on their branch after phastSim, resulting in their collapse with their parent. "Parsimony" is the total tree parsimony score, or the count of all mutations across all branches, and also reflects mutations per node and scale. "Tree depth" is the mean distance in mutations between the root of the tree and a leaf. ARI, or adjusted rand index, is the computed adjusted rand index for sample cluster labels on the final collapsed tree versus the true clusters. True clusters here are defined as the set of samples which share a single true migration event into their region at their common ancestor. IAC stands for "internal assignments correct", or the proportion of internal nodes which have their true regional states correctly assigned by the heuristic on the uncollapsed, bifurcating tree.

| Sample Count | Samples In-Region | Time (seconds) |
|---|---|---|
| 100 | 25 | 0.02106654644 |

| | | |
|---|---|---|
| 1000 | 250 | 0.03384798765 |
| 10000 | 2500 | 0.2106143832 |
| 50000 | 12500 | 0.9356530309 |
| 100000 | 25000 | 1.890872002 |
| 500000 | 125000 | 9.770167351 |
| 1000000 | 250000 | 19.75107902 |
| 2500000 | 625000 | 37.4885782 |

**Supplementary Table 2:** Basic benchmarking information for our method. For this benchmark, we took the public tree obtained on 11-08-21 and randomly generated subtrees containing a set number of samples. We further selected at random 25% of these samples to be considered in-region, under a single region model. We find that runtime is approximately linear with the number of samples in the tree (which, in turn, is correlated with the number of nodes in the tree). Even for a tree of two and a half million samples and a region with 625,000 samples, a single region on a single thread doesn't take more than one minute to compute our heuristic for.

**Supplementary Table 3:** Inferred introduction counts to each of the fifty United States from international sources.

**Supplementary Table 4:** Inferred introduction counts between each of the fifty United States.
